# DACr - An Algorithm for Treating Diverse Missing Values in Large Data, with Application to Heart Transplantation

**DOI:** 10.64898/2026.01.20.26343799

**Authors:** Wei Dai, Jiayang Sun, Jie Xu

## Abstract

The increasing use of Real-World Data (RWD) in clinical research is critical for evidence-based decision making but presents challenges to data analytics. Unlike in Randomized Controlled Trials (RCTs), missingness in RWD occurs often and can include complex patterns which may or may not be Missing at Random (MAR). Informative absence (Missing Not at Random, or MNAR) occurs when the absence of data itself is a clinical signal. Applications of ad-hoc methods, or even popular “universal” methods, can lead to biased inferences when these varied data problems are not appropriately addressed. This paper introduces a multi-step algorithm known as **D**ivide **A**nd **C**onquer, **r**ejoin (DACr, pronounced as “DACK-er”) for handling missing data in RWD. We illustrate the algorithm’s application using a large-scale, high-dimensional heart transplant dataset (from *p* = 1107 to 527 after initial screening) from the Scientific Registry of Transplant Recipients (SRTR). This “divide and conquer” idea handles variables based on data type, the proportion, and type of missingness. It also allows for the targeted application of justified data handling techniques: appropriate imputation method is used for variables assumed to be MAR, while variables with plausible MNAR mechanisms are recoded as categorical with an extra level of “missing” to preserve their informative signal. The proposed algorithm provides a systematic treatment for practical researchers.

## 1 Introduction

Mining clinical evidence is increasingly reliant on Real-World Data (RWD), such as data from Electronic Health Records (EHRs), patient registries, and administrative claims [1]. Unlike data from rigidly controlled Randomized Controlled Trials (RCTs), RWD is “found data,” [2] collected retrospectively for clinical, administrative, or billing purposes, initially not necessarily for research. This difference in data-generating processes presents a gap, particularly in the handling of incomplete data. In the controlled environment of RCTs, the mechanisms underlying missing data are more clearly understood and can therefore be addressed prospectively through formal frameworks. The ICH E9 (R1) addendum [3] on estimands and intercurrent events (ICE), for example, forces researchers to pre-specify the precise scientific question and how events like treatment discontinuation or rescue medication use will be handled. This paradigm shifts the discussion from a reactive statistical “clean-up” to a proactive definition of the causal question [4].

This formal, prospective framework is, however, incompatible with RWD. In observational data, missingness may not be a protocol deviation but an inherent, complex feature of the data itself. The mechanisms driving missingness are varied: data may be Missing Completely at Random (MCAR) due to an equipment failure; Missing at Random (MAR), where missingness is predictable from other observed variables; or, most critically, Missing Not at Random (MNAR). In RWD, MNAR is common and often takes the form of informative absence. For example, a clinician may not order a lab test because the patient appears healthy, or a follow-up visit is missing because the patient’s condition worsened [5], [6]. In these scenarios, the absence of data is itself an informative signal. Applying naive or ad-hoc solutions to this complex problem, such as listwise deletion (or Complete Case Analysis, CCA) or single imputation without curating the data can lead to biased inferences [6]. Listwise deletion is valid only under the strong and rare MCAR assumption [7], while single imputation methods artificially reduce variance, distort associations, and can lead to conclusions that are the opposite of the truth [8]. Even principled methods like multiple imputation (MI), which are valid under MAR, will produce biased results if applied naively without considering the plausible MNAR [5]. Therefore, RWD requires a systematic algorithm where each data-handling decision is justified by a plausible assumption about its underlying missingness mechanism and clinical context. Literature has established guidelines for the systematic curation of RWD to ensure statistical validity. The EDPAI framework [9] formalizes the conversion of heterogeneous EHR entries into structured datasets through a taxonomy of extraction, feature definition, and value assessment. Their work aligns with the FDA’s 2024 Guidance[10], which mandates a transparent provenance chain to substantiate data fitness through the dual criteria of reliability and relevance. While these works provide essential high-level guidance, this paper introduces the DACr algorithm to provide a granular implementation. The algorithm is presented not as a universal solution, but as a sensible and practical template for moving beyond simplistic fixes. We apply this algorithm to a large-scale, high-dimensionality heart transplant dataset from the Scientific Registry of Transplant Recipients (SRTR) to illustrate its clinical application.

## 2 Review

Rubin [11] provided a foundational taxonomy that classifies missing data mechanisms into three distinct categories: Missing Completely at Random (MCAR), Missing at Random (MAR), and Missing Not at Random (MNAR). The choice of an appropriate statistical method for handling missing data depends on which of these mechanisms is assumed to be in operation, and also their percentages of missingness.

We adopt a commonly accepted notation for missing values. Let *Y*_*i*_ be a variable of interest for a subject *i*, which may be unobserved. Let *X*_*i*_ be a vector of other observed variables (e.g., baseline covariates) for the same subject. We define a missingness indicator, *δ*_*i*_ for *Y*_*i*_ such that,

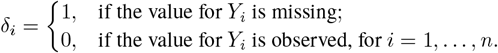

The full data for this context consists of *n* observations, (*Y*_*i*_, *X*_*i*_), *i* = 1, …, *n*. The observed data consists of 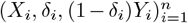. The missing data mechanism is characterized by the conditional probability of missingness for *Y*_*i*_, given the full data, ℙ(*δ*_*i*_ = 1 | *X*_*i*_, *Y*_*i*_) for *i* = 1, …, *n*.

### Missing Completely at Random (MCAR)

The MCAR mechanism assumes that the probability of missingness is independent of all variables, whether observed or unobserved. This is the simplest and most restrictive assumption. It implies that the missingness is a purely random process. Statistically, the probability of missingness does not depend on either *X*_*i*_ or *Y*_*i*_,

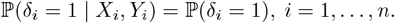

Under MCAR, the observed values of *Y* are a simple random sample of the complete set of *Y* values. Consequently, using a listwise deletion will produce unbiased estimates of the mean or regression coefficients although it may significantly decrease remaining number of observations.

### Missing at Random (MAR)

The MAR mechanism is a more relaxed and often more plausible assumption. Under MAR, the probability of missingness is assumed to depend *only* on the observed data (*X*_*i*_), and not on the unobserved data (*Y*_*i*_) after conditioning on the observed data. This implies that, conditional on the observed information, the missingness is random. Statistically, the probability of missingness is independent of the missing value *Y*_*i*_ once we condition on the observed variables *X*_*i*_,

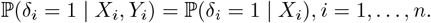

The MAR assumption is vital to many missing data methods, such as multiple imputation.

### Missing Not at Random (MNAR)

The MNAR mechanism, also known as non-ignorable missingness, is the most problematic case that cannot assume MCAR or MAR. It occurs when the probability of missingness depends on the value of the missing variable *Y*_*i*_ itself, even after conditioning on the observed data *X*_*i*_. This mechanism is “non-ignorable” because the missingness process is explicitly linked to the unobserved value. Formally, the conditional probability cannot be simplified and remains dependent on *Y*_*i*_,

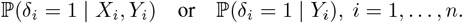

MNAR is the most challenging scenario because the missing data mechanism must be explicitly modeled as part of the statistical analysis to avoid biased outcome. Sometimes, a common practice in the MNAR case, mentioned in Fang [12], is to “*conduct sensitivity analysis to explore the robustness of the main results under the MNAR assumption*.*”* Practitioners should consider biased sampling models and recent developments subject to data shifts [13], such as those regarding treatment as advocated below, to incorporate informative missingness as a distinct level in the data analysis.

## 3 Method

In this section, we detail the proposed DACr, a stratified preprocessing algorithm designed to handle high-dimensional datasets with heterogeneous missingness mechanisms.

### 3.1 DACr Algorithm

The algorithm workflow is illustrated in Figure 1. Before the implementation of the proposed algorithm, it is assumed that a comprehensive exploratory data analysis (EDA) has been conducted to characterize the underlying dataset. The procedure begins with the exclusion of irrelevant predictors, guided by the `clinical_inputs` argument to ensure that domain-specific expertise and essential clinical surrogates are preserved prior to further data preprocessing. The algorithm then **divides** the remaining variable space into two distinct subsets, represented by `x_cat` for categorical data and `x_num` for numerical data.

**Figure 1.**
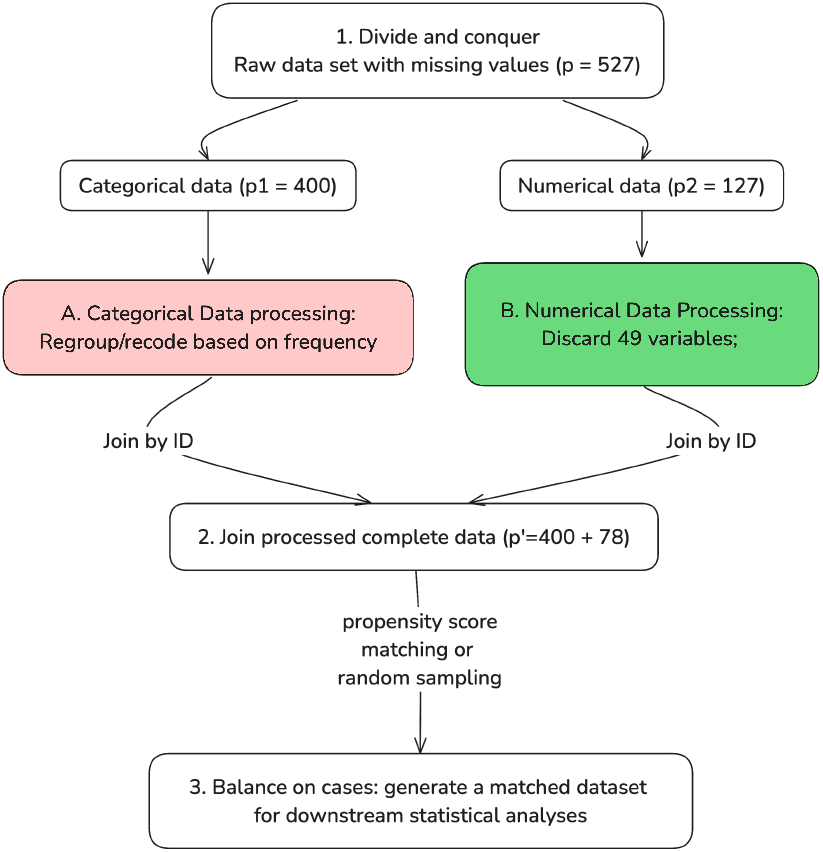
the Divide and Conquer, rejoin Algorithm

After partitioning, we analyze whether missing data patterns in each covariate relates to the response variable to determine if the missingness is informative. The most intuitive method to determine if missing data patterns were informative is through a visualization. We can also utilize decision tree stumps by determining if a single split based on missingness reduced impurity. Additionally, we can apply formal hypothesis testing to confirm statistical significance between the missing data and the outcome.

The extent of missing data is quantified and stratified based on `thresholds` informed by the EDA, typically distinguishing between low, medium, and high proportions of missingness. These `thresholds` serve as decision boundaries for the data processing pipeline. For example, a low threshold might identify variables with minimal data loss, while a high threshold identifies variables where the degree of missingness may become unacceptable and compromise the stability of the statistical estimates.

For variables within `x_cat`, if the missingness is found to be informative or relevant to the outcome, an additional level is introduced to the variable to explicitly handle the missing state. Conversely, if the missingness lacks predictive relevance, the variable is either removed from the analysis or addressed via the strategies specified in the `mechanisms` argument, such as MI.

For the remaining relevant covariates in `x_num`, the treatment is determined by the assumed nature of the missingness mechanism. Variables identified as MNAR are recoded as categorical variables to preserve the systematic information contained within the missing values. For variables adhering to the MAR assumption, the treatment is stratified by the proportion of missingness: variables with high missingness are typically removed to maintain model parsimony, those with medium missingness are addressed through MI, and those with low missingness are managed either via MI or by removing the incomplete cases. So far, we have **conquered** missing values in categorical and numerical variables, the most common types found in tabular RWD.

Finally, the processed subsets are **rejoined** to form the complete dataset. This completes the DACr algorithm. Depending on the research question and the distribution of the response variable, balancing strategies may be optionally implemented to address potential outcome imbalances.

Conceptually, the algorithm can be summarized as a high-level function with inputs:

~~~
DACr(
  x_cat, x_num,
  clinical_inputs,
  mechanisms,
  thresholds = c(0.01, 0.5),
  …).
~~~

## 4 Results on SRTR Data

We illustrate with the study utilized observational data from the Scientific Registry of Transplant Recipients (SRTR). The SRTR dataset is a national-level registry containing detailed pre-operative, operative, and post-operative data on organ transplant recipients in the United States. As with many large-scale, real-world observational datasets, the raw data (*p* = 527 variables after initial screening) is characterized by high dimensionality and a heterogeneous missing data problem.

### 4.1 Data Preprocess

The missing data in the SRTR dataset is complex because its primary purpose is to collect administrative and clinical data from multiple centers over time. Data may be missing for various reasons, many of which have plausible explanations that fall into Rubin [11]’s categorization. For instance, we identified one observation with a BMI value greater than 1000. This is clearly an erroneous data entry, which can be reasonably assumed to be MCAR, as the error is likely independent of other patient characteristics. Other variables, such as “recipient ischemic time,” are plausibly MAR, where missingness may be predictable from other observed variables. Finally, a lab test value that is missing because the test was not performed represents a clinically informative absence. This may indicate the patient appeared healthy to some extend or, conversely, was unsuitable for the test. In either case, the missingness is related to the unobserved clinical state itself and is therefore MNAR. Given this complexity, simple ad-hoc methods like listwise deletion or mean imputation would introduce unacceptable bias and loss of statistical power. Therefore, we applied DACr algorithm to preprocess the data, handle missing values and prepare a balanced cohort for downstream analysis.

### 4.2 Results

The dataset was first stratified by variable type, yielding two distinct subsets: *p*_1_ = 400 categorical variables and *p*_2_ = 127 numerical variables. This stratification facilitated the use of type-specific processing. For the categorical subset, variables were processed by regrouping or recoding infrequent levels.

A more complex approach was required for the *p*_2_ = 127 numerical variables, which exhibited heterogeneous missingness as illustrated in Figure 2. These variables were partitioned into three strata based on the proportion of missing values: high missingness (>50%), moderate missingness (1%−50%), and low missingness (<1%). It is important to note that the choice of such strata is data-specific. After examining the data missingness, we determined that these strata best facilitate the treatment of each subset.

**Figure 2.**
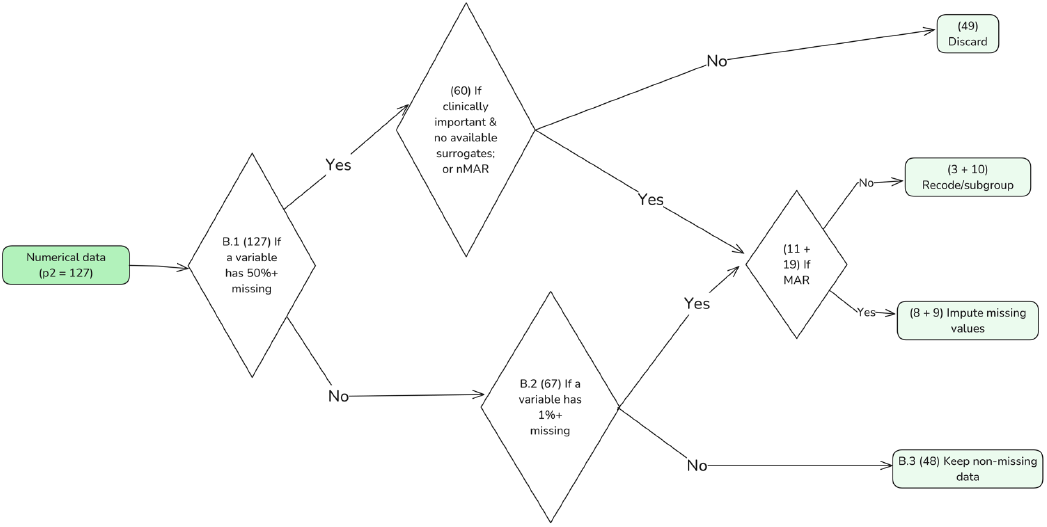
the DACr Algorithm Workflow for Numeric Data

For the high missingness stratum (*n* = 60), variables were first subjected to a joint clinical and statistical review to assess their importance and redundancy. This expert-led review resulted in the exclusion of *n* = 49 variables deemed non-essential for the research question or for which a high-quality surrogate was available, thereby reducing model dimensionality. The remaining *n* = 11 clinically important variables were handled based on their assumed missingness mechanism. For *n* = 8 variables plausibly assumed to be MAR, a regression-based Multiple Imputation technique was employed. Conversely, *n* = 3 variables were determined to be MNAR, where the absence of a value was itself informative. To preserve this information without introducing bias, these variables were recoded as categorical, creating a distinct “Missing” level to allow the model to learn from the missingness pattern.

A similar approach was applied to the moderate missingness stratum (*n* = 19), although we skipped the domain expert review at this stratum. For the *n* = 9 variables assumed to be MAR, Multiple Imputation was performed. The other *n* = 10 variables, assumed to be MNAR, were likewise categorized with a “missing” level to retain the informative nature of their absence.

Finally, for variables in the low-missingness stratum (*n* = 48), data were assumed to be MCAR. While imputation is often preferred to maximize statistical power, the proportion of missing data in this stratum was trivial. Consequently, listwise deletion was employed. In our case, around only 3% of observations were removed at this step, which has a negligible impact on statistical power or bias.

Following these processing steps, the processed categorical (*p*_1_ = 400) and the processed numerical (*n* = 78) datasets were joined by a unique patient identifier to form the final, complete dataset for analysis. A concise workflow is presented in Figure 3.

**Figure 3.**
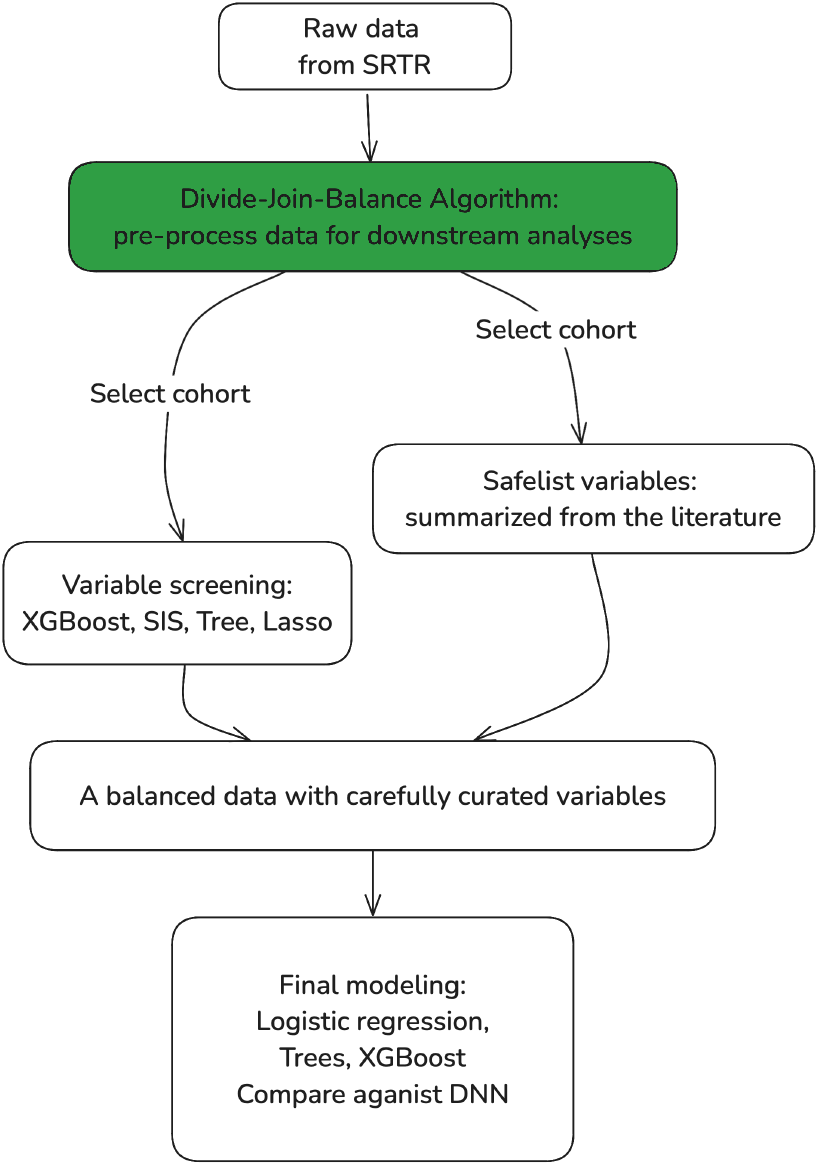
the Complete Data Analysis Illustration

## 5 Discussion

The DACr algorithm provides a systematic framework for handling the heterogeneous missing data to RWD. While this stratification enhances the plausibility of the applied methods, we acknowledge that the choice of missingness thresholds (e.g., 1% and 50%) is inherently data-specific and introduces a degree of subjectivity.

A further consideration not addressed in this pipeline, particularly for downstream predictive modeling, is the prevention of data leakage between training and testing datasets. Data leakage occurs when information from outside the training dataset is improperly used to inform the model, a common issue in complex preprocessing that leads to an overly optimistic estimation of model performance [14]. For example, if a variable is imputed using a median derived from the entire dataset before partitioning, information from the test set has “contaminated” the training set. The missing data handling methods must be based only on the training data, ensuring the test set remains a true, independent evaluation of the pipeline’s generalizability. This principle applies to all data-handling steps, from calculating medians to fitting complex imputation models.

## Data Availability

Data for this study were provided by the Scientific Registry of Transplant Recipients (SRTR) and are not available for the general public due to data use restrictions. Researchers may apply for access to these data directly through the SRTR at www.srtr.org.

